# The Effects of Nurse Specialty Certification and Medical Treatments on Glycemic Control Outcomes: A Retrospective Study

**DOI:** 10.1101/2020.05.26.20113936

**Authors:** Maria M. Ojeda

**Author notes:** Authorship: M.M.O. designed the study, collected the data, analyzed the data, and prepared this manuscript.

## Abstract

**AIMS:** To describe and explore the impact of the presence of specialty certified nurses on the unit on glycemic control among non-critically ill hospitalized patients with Diabetes mellitus.

**BACKGROUND:** Poor glycemic control during hospitalization has been associated adverse patient outcomes. Staff nurses administer much of the diabetes-specific care delivered to patients during hospitalization. Nurse specialty certification is a recognized indicator of nursing care quality.

**METHODS:** A retrospective sample of medical records data was analyzed to describe and explore the impact of specialty certified nurses on glycemic control among a group of 293 non-critically ill hospitalized patients with Diabetes mellitus. Regression analysis was performed to assess the impact of the percentage of certified nurses on the unit as a mediator or moderator of the relationship between medical treatments and glycemic control outcomes.

**RESULTS:** Increases in the percentage of specialty certified nurses on the unit were associated with significant decreases in mean blood glucose levels among patients who received a basal-bolus insulin regimen or enteral nutritional feedings.

**CONCLUSIONS:** Patients who received specific medical treatments during hospitalization had superior outcomes when nursing care was delivered during times in which the relative proportion of certified nurses on the unit were higher.

## Introduction

The prevalence of diabetes in the United States is high, and continues to grow. People with diabetes are disproportionately represented among those admitted to acute care hospitals. In 2012, 25.7% of all inpatient hospital days and approximately $174 billion (26%) of hospital expenditures were attributed to diabetes (American Diabetes Association [ADA], 2013). Glycemic control (GC) may be defined as the avoidance of extremes in blood glucose levels. In hospitals, poor GC is an important public health problem that has strong economic and patient safety implications, due to its association with poor patient outcomes. Poor GC during hospitalization is associated with increased hospital length of stay, transfers to higher levels of care, increased incidence of nosocomial infections, poor functional outcomes, and increased mortality rates among patients with diabetes (Brock, Shirley, Bardgett, Walker, & Deehan, 2017; Draznin et al., 2014; Falciglia, Freyberg, Almenoff, Alessio, & Render, 2009; Frisch et al., 2010; Olveira et al., 2013; Subramaniam et al., 2014; Williams et al., 2017). While there is convincing evidence that improving GC during hospitalization for acute illness can effectively reduce morbidity and mortality and improve outcomes, poor GC continues to be a pervasive problem among non-critically ill hospitalized patients who have diabetes (Arnold, Mahesri, McDonnell, & Alexanian, 2017; Cook et al., 2007; Cook et al., 2009; Mendez et al., 2013; Murad et al., 2012; Swanson, Potter, Kongable, & Cook, 2011; Vogel, Smith, & Kruse, 2017).

Medical treatments, both related and unrelated to the treatment of diabetes have been identified as factors that differentially contribute to GC in non-critically ill hospitalized patients. The amount and type of medications prescribed for the inpatient treatment of diabetes (e.g., insulin and oral diabetes medications), and commonly prescribed treatments and medications used in the care of conditions other than diabetes, such as beta-blockers, glucocorticoids, and enteral and parenteral nutrition are associated with variations in hypoglycemia and hyperglycemia rates during hospitalization (Dickerson, Ye, Sack, & Hueston, 2003; Khan, Golden, Mathioudakis, 2017; Lin et al., 2010; Llop et al., 2012; Olveira et al., 2013; Umpierrez et al., 2007; Umpierrez et al., 2011; Varghese et al., 2007; Vindedzis, Marsh, Sherriff, & Stanton, 2014). In addition to specific medical treatments, the quality of diabetes-specific care delivered to hospitalized patients might also affect GC. Experts have often cited nursing care as key in achieving GC in hospitalized patients (ADA, 2015; Moghissi et al., 2009; Sanson, Vellone, Kangasniemi, Alvaro, & D’Agostino, 2017). Diabetes-specific nursing care is a ubiquitous and complex aspect of inpatient nursing practice, involving multiple components that must be accurately performed and coordinated in a timely manner to maintain the quality and safety of care delivered (Freeland, Penprace, & Anthony, 2011; Klinkner, 2016; Malesker et al., 2007). Nonetheless, little is known about how acute care hospital nursing impacts outcomes attributable to GC.

Researchers of general inpatient populations have identified nursing staff characteristics (staffing levels, specialty certification rates) as indicators of inpatient nursing care quality associated with improvements in patient outcomes such as mortality, nosocomial infections, falls, and readmission rates (Aiken et al., 2010; Aiken et al., 2014; Boltz, Capezuti, Wagner, Rosenberg, & Secic, 2013; Boyle, Cramer, & Potter, 2014; Kane, Shamliyan, Mueller, Duval, & Wilt, 2007; Kendall-Gallagher, Aiken, Sloane, & Cimiotti, 2011; Kendall-Gallagher & Blegen, 2009; Krapohl, Manojlovich, Redman, & Zhang, 2010; McHugh, Berez, & Small, 2013; Needleman et al., 2006; Tourangeau et al., 2007). Nonetheless, there is a scarcity of research examining the relationship between indicators of inpatient nursing care quality and GC in acute care hospitals. Only one published study has investigated the relationship between nurse staffing levels and GC, however that study did not examine the impact of staff nurses with a specialty certification on inpatient GC (McHugh, Shang, Sloane, & Aiken, 2011).

### Background

Despite continued efforts by professional nursing organizations and employers to encourage specialty certification among nursing staff (Academy of Medical-Surgical Nurses [AMSN], 2016; American Association of Critical-Care Nurses [AACN], no date; American Board of Nursing Specialties [ABNS], 2005; American Nurses Credentialing Center [ANCC], 2012; Williams, Lopez, & Lewis, 2013), little is known about the effects of nursing certification on patient outcomes. Only, four published studies addressing the effect of nurse certification on outcomes in hospitals and among acute care hospital inpatient populations were found. The first study examined the relationship of certification among nurses working in intensive care units (ICU) to the occurrence of adverse events (Kendall-Gallagher & Blegen, 2009). The retrospective correlational study involved a secondary analysis of unit-level data. Twenty-nine randomly selected hospitals and 48 adult ICUs (31 medical-surgical ICUs and 17 cardiac ICUs) were included in the study. The relationship of the proportion of certified nurses on each unit to the occurrence of medication administration errors, patient falls, skin breakdown, and nosocomial infections was investigated. In multivariate analysis using hierarchical regression modeling controlled for hospital-level characteristics, the higher the percentage of certified nurses on the unit, the lower the frequency of patient falls (*p* = .04). Nurse certification was not associated with rates of medication errors, skin breakdown or nosocomial infections. The second study involved the secondary analysis of 450 survey responses; surveys were collected from staff nurses on 25 intensive care units, in 8 hospitals in Michigan (Krapohl et al., 2010). The purpose was to determine whether the proportion of certified nurses mediated the relationship between nurses’ workplace empowerment and the rate of adverse outcomes within acute care hospital units. Rates of certification were found to vary widely between individual units, the mean rate of certification for the overall sample was 17%. Nurses’ perceptions of workplace empowerment were positively associated with the proportion of certified nurses per unit (*p* = .05), however nurse certification rates were not associated with any patient outcomes. Later, Boltz and colleagues (2013) investigated the relationship between nurse certification and indicators of patient safety. The retrospective descriptive study was conducted using data from 44 medical-surgical units at 25 different hospitals. The researchers found that units with high proportions of nurses with any specialty certification (> 5%) had significantly lower fall rates (*p* = .05) but no association was found to falls with injuries, pressure ulcer prevalence, or the prevalence of the use of physical restraints. The most recent study examined the relationship of specific types of nurse certifications and outcomes among post-surgical patients (Boyle, Cramer, & Potter, 2014). A total of 178 surgical ICUs and 269 surgical units were included in the study. The study design was a secondary data analysis of the National Database of Nursing Quality Indicators (NDNQI) database for the year 2011. The percentage of nurses holding certification as a certified ambulatory perianesthesia nurse (CAPA), certified postanesthesia nurse (CPAN), certified operating room nurse (CNOR), or certified registered nurse first assistant (CRNFA) was examined, as well as the percentage of nurses holding any type of certification. Quarterly data on certification was used to predict NDNQI-listed patient outcomes the following quarter using generalized linear modeling. Mean rates of nurse certification within each unit ranged from 14.1% to 63%. In regression models adjusted for hospital (Magnet® status, bed size, teaching status, hospital case-mix index) and nursing staff characteristics (staffing levels, years of experience, proportion of nurses with a baccalaureate degree, practice environment), a 10% increase in CPAN certification rates, decreased CLABSI by 8% (*p* = .05) and a 10% increase in CNOR and CRNFA certification was associated with a 16% decrease in CLABSI (*p* = .00) among patients in surgical ICUs the following quarter. In non-ICU surgical units, a 10% increase in the rates of CNOR and CRNFA certification were associated with an 8% increase in hospital-acquired pressure ulcers (*p* = .03) and a 14% increase in unit-acquired pressure ulcers (*p* < .01) the following quarter.

Taken together, the results of these studies warrant further investigation of the association between nurse certification and patient outcomes (Boltz et al., 2013; Boyle et al., 2014; Kendall-Gallagher & Blegen, 2009; Krapohl et al., 2010). In 3 of 4 studies, an increase in nurse certification was associated with select patient outcomes including falls and rates of CLABSI (Boltz et al., 2013; Boyle et al., 2014; Kendall-Gallagher & Blegen, 2009). Nonetheless, Boyle and colleagues (2014) found that an increase in nurse certification rates was associated with an increase in rates of hospital and unit-acquired pressure ulcers among non-critically ill surgical patients; an earlier study (Boltz et al., 2013) obtained mixed results, and another study found no association between nurse certification and patient outcomes (Krapohl et al., 2010). None of those studies controlled for potential differences in the medical treatments administered to patients that may have contributed to the observed results.

In summary, little is known about the effects of nurse certification on patient outcomes among non-critically ill patients. Studies examining the effects of nurse certification on patient outcomes in the context of specific medical treatments administered are missing from the research literature and there have been no studies to date have described the relationship of nurse certification to inpatient GC.

### Purpose

The purpose of this study was to examine whether nurse certification impacted GC among non-critically ill patients with diabetes who were hospitalized for any cause. The study explored the potential effects of nurse certification on GC in the presence of specific medical treatments. To further inform our understanding of this potentially complex relationship, the association between nurse certification and nurse staffing was also examined.

#### Research question

The research question the study sought to answer was: Does the percentage of certified nurses, mediate or moderate the effects of medical treatments (glucocorticoid medications, beta-adrenergic blockers, oral antidiabetes medications, type of insulin regimen, and nutritional feedings) on GC (mean blood glucose level, combined percentage of hypoglycemic and hypoglycemic blood glucose testing results, and glycemic variability) among non-critically ill hospitalized patients with diabetes mellitus?

## Methods

Retrospective data was used to describe and explore the relationship between nurse certification, specific medical treatments administered, and GC among non-critically ill patients hospitalized with diabetes. Ethical approval from the Institutional Review Board (IRB) was obtained for the study in December of 2015. The study sample was limited to non-critically ill hospitalized patients admitted to one of four medical-surgical units. A list of all patient records with a diagnosis of Type-1 or Type-2 diabetes (based on ICD-9 codes) for the study period (January 2014 – June 2014) was created from one of the study hospital’s administrative databases. Inclusion criteria were 1) a primary or secondary admission diagnosis of Type-1 or Type-2 diabetes, 2) age of 18 years or older and receiving point-of-care blood glucose testing during their hospital stay, and 3) hospital length of stay (LOS) of ≥ 1 day. Cases were excluded if 1) the patient had initially been admitted to a non-medical-surgical unit (e.g., intensive or progressive care, labor and delivery, pediatric units), or 2) the patient had been admitted for extreme manifestations of poor GC (ketoacidosis, hyperosmolar coma, hypoglycemic coma). Due to the exploratory nature of the study, the minimum sample size of hospital admission records included in this study was *N* = 270, based on the *N:q* rule (Kline, 2011). Oversampling to account for the possibility of missing data was used, and thus from the pool of patient records that met eligibility requirements, a total of 300 patient records were randomly selected for analysis.

### Study Variables

Variables representing patient characteristics were included for descriptive purposes only. Medical treatments, nurse certification, nurse staffing, and GC were measured at the patient level in order to reflect real-world conditions at the time period in which individual patients were hospitalized. Data on unit-specific nurse certification were linked to each individual patient’s record to reflect the actual period of hospitalization.

#### Patient characteristics

Demographic patient characteristics were gathered from patients’ medical records and included age (in years), gender (Male, Female), race/ethnicity (White non-Hispanic, White-Hispanic, Black, Asian/Pacific Islander, Other). Variables representing clinical patient characteristics included type of diabetes (Type-1, Type-2), serum glycated hemoglobin level on hospital admission, comorbidities, and indicators of diabetes-specific severity of illness. The Charlson Comorbidity Index (CCI) was used to measure patient comorbidities (Charlson et al., 1987). The CCI is a weighted index of patient comorbidities based upon associated risk for short and long-term (10-year) mortality. Like previous studies focusing on patients with diabetes this study used a modified version of the CCI that excluded the scoring of diabetes in order to explore how other comorbidities may have contributed to GC and patient outcomes; the range of possible scores was between zero and thirty (Holman, Hillson, & Young, 2013; Turchin et al., 2009). The Diabetes Complications Severity Index (DCSI) was used to measure the impact of diabetes-related severity of illness. The DCSI was originally developed to predict mortality and risk for hospitalization based upon the presence and severity of diabetes-related complications and serum creatinine levels (Young et al., 2008). The DCSI classifies diabetes-complications into seven categories and 55 individual conditions with corresponding ICD-9 codes. This study used an adapted version (aDCSI) which excludes serum creatinine testing data; the range of possible scores on the aDCSI was zero and thirteen (Chang, Weiner, Richards, Bleich, & Segal, 2012; Chang, Weiner, Richards, Bleich, & Segal, 2012a). Data collected for scoring each index (i.e., CCI & aDCSI) were extracted from hospital administrative databases using ICD-9 codes.

#### Medical treatments

Medical treatments were defined as medically prescribed treatments that may modify the risk for poor GC during hospitalization. Specific treatments were chosen based on the extant evidence, they consisted of treatments targeting glycemic management during the hospitalization period (e.g., insulin therapy, oral antidiabetes medications), as well as other treatments that had the potential for modifying the risk for poor GC (e.g., nutritional feedings, beta-adrenergic blockers, and glucocorticoids). Variables were coded to represent whether the medication had been administered to the patient during hospitalization. The variables representing oral antidiabetes medications, beta-adrenergic blockers, glucocorticoids, and nutritional feedings (i.e., enteral or parenteral) were dichotomous in nature (not administered, administered). The variable representing insulin was represented categorically (no insulin, sliding-scale, or basal-bolus insulin).

#### Nurse certification

Nurse certification was defined as a nationally-recognized professional specialty certification of any type. Nurse certification was measured as the percentage of nurses with a specialty certification on the unit during the period of the patient’ s hospitalization.

#### Nurse staffing

Two measures were used to represent nurse staffing levels: total RN hours per patient day and registered nurse (RN)-to-patient ratios, as available within the organization’s administrative database. Each daily measure of nurse staffing was aggregated at the patient level as an average on the unit for the entire length-of-stay.

#### Glycemic control

Glycemic control was defined using the ADA’s (2015) standards of practice for non-critically ill hospitalized patients. Random blood glucose levels of 70-180 mg/dl were considered euglycemic; dysglycemia was defined as the cumulative percentage of blood glucose levels reflecting hypoglycemia (<70 mg/dl) and hyperglycemia (>180 mg/dl). A secondary aspect of GC is glycemic variability. Glycemic variability is associated with increased LOS, inpatient complications, and mortality (Draznin et al., 2014; Mendez et al., 2013). Glycemic variability in this study was measured as the standard deviation of the mean blood glucose level for each patient, it is defined as the magnitude of the variation in blood glucose levels within individual patients throughout their hospital stay.

#### Data Collection Procedures

Patient data was retrospectively collected using a combination of hospital administrative data, medical record review data, and laboratory reports of point-of-care blood glucose testing data. Self-reported data regarding the proportion of nurses with specialty certification within each unit were included in this study, such data are routinely collected on a quarterly basis by hospital administration and stored in a unique database. Reviews of each medical record included in the study was conducted to determine whether specific medical treatments that might affect GC were administered during hospitalization. Glycemic control was derived from data stored in a clinical laboratory database and consisted of point-of-care blood glucose testing results for each patient beginning with the 2^nd^-day of hospitalization.

### Analyses

All analyses were conducted using IBM SPSS V. 23, with add-on feature PROCESS V. 2.13.2 a level of significance of *α*=0.05 used for all analyses (Hayes, 2015; IBM Corporation, 2015). Data screening and preparation was conducted by examining the dataset for missing elements, multivariate outliers, and for the underlying assumptions of multivariate analysis prior to analysis. Whenever possible, data transformations were performed to improve approximation of a normal distribution. Descriptive statistics were generated using frequencies, percentiles, and measures of central tendency. Spearman’s *rho* was used to examine potential relationships between nurse certification and the variables representing nurse staffing (total RN hours per patient day and RN-to-patient ratios).

Mediators are variables that explain the relationship between a predictor and an outcome (Jose, 2013). Ordinary least squares regression analysis was conducted separately for each medical treatment to explore the role of nurse certification in the causal pathway between each medical treatment and GC. A significant indirect effect (Hayes, 2013) indicated that nurse certification could explain the relationship between medical treatments (predictor) on GC (outcome). Moderators are variables that affect the strength and direction of the relationship between predictors and outcomes (Jose, 2013). Hierarchical multiple regression was used to conduct moderation analysis. Separate models were generated for each medical treatment and the predictive value of each medical treatment and nurse certification on GC was assessed. An interaction term consisting of a medical treatment and nurse certification was added to each of the models. A significant interaction (significant *F*-test and significant increase in *R*^2^, along with a bootstrap confidence interval that did not contain “0” [Hayes, 2013]) indicated that the nurse certification influenced the nature of the relationship between medical treatments and GC.

## Results

For the 6-month study period in 2014, a total of 1,706 medical records of patients hospitalized on four medical-surgical units were eligible for inclusion in the study; 300 of those records were randomly selected for inclusion in the analysis. Six records were found to be multivariate outliers (Critical Value: *χ^2^* = 32.90, *df* = 12, *p <* .001) and one record did not meet the age restrictions of the study and were removed. The total number of records included in the study was 293. There was no missing data among variables included in regression modeling. However, data for the variable representing glycated hemoglobin level on admission was missing from 68 of the records, descriptive statistics for this variable are reported based on a reduced sample.

### Description of the Sample

The average age of patients included in the sample was 64.5 years (*SD* = 14.7, *N* = 293). The majority of the patients were female, had Type-2 diabetes, and were White-Hispanic; most patients, had at least one additional chronic condition and one or more diabetes-related complications documented within the medical record (Table 1). The average glycated hemoglobin level on admission was 7.9% (*n* = 225), greater than the ADA’s (2015) recommended goal of < 7%. The mean point-of-care blood glucose level recorded for the overall sample of 293 patient records was 162 mg/dl, and 30% (*n* = 88) had at least half of the recorded blood glucose testing results that indicated dysglycemia (hypoglycemia or hyperglycemia) or had an average variation of less at least 50 mg/dl between each blood glucose testing result. Fifty-eight (19.8%) patients received glucocorticoid medications, 113 (38.6%) patients received beta-adrenergic blocker drugs, and 16 (5.5%) of patients received oral antidiabetes medications while hospitalized. The majority of patients, 168 (57.3%), received a basal-bolus insulin regimen for GC during their hospital stay. Ninety-five (32.4%) of patients received sliding-scale insulin, while 30 (10.2%) patients did not receive any insulin as part of their inpatient treatment regimen. Eleven patients (3.8%) were administered nutritional feedings during hospitalization; all of the feedings were via the enteral route.

**Table 1.** Description of Patient Characteristics (*N* = 293).

| Characteristic | *n* | % |
| --- | --- | --- |
| Gender |  |  |
| Female | 159 | 54.3 |
| Male | 134 | 45.7 |
| Ethnicity<sup>†</sup> |  |  |
| White | 64 | 21.8 |
| White – Hispanic | 130 | 44.4 |
| Black | 96 | 32.8 |
| Asian/Pacific Islander | 2 | 0.7 |
| Other | 1 | 0.3 |
| Type of Diabetes |  |  |
| Type-1 | 5 | 1.7 |
| Type-2 | 288 | 98.3 |
| Charlson Comorbidity Index Score |  |  |
| 0 | 78 | 26.6 |
| 1 | 55 | 18.8 |
| 2<sup>††</sup> | 55 | 18.8 |
| 3 | 34 | 11.6 |
| 4 | 33 | 11.3 |
| 5 or greater | 38 | 12.9 |
| Adapted Diabetes Complications Severity Index Score |  |  |
| 0 | 77 | 26.3 |
| 1 | 52 | 17.7 |
| 2<sup>††</sup> | 73 | 24.9 |
| 3 | 29 | 9.9 |
| 4 or greater | 62 | 21.2 |

*Note.* <sup>†</sup>Black-Hispanic ( $n = 6$ ) grouped with Black; no Native Americans/Alaskan
Natives included in the sample. <sup>††</sup>Median.

During the study period, the median RN hours per patient day was 117.3 (*range* = 159.2), the median RN-to-patient ratio was 5.6 (*range* = 13.9). The median percentage of nurses with a specialty certification was 34.8%; 10.6% (*n*= 31) of patients were hospitalized during periods in which the percentage of specialty certified nurses on the unit was lowest (< 25%), 52.9% (*n*= 155) when the percentage was moderate (25% - 35%) and the remaining 36.5% (*n*= 107) when the percentage was highest (> 35%). There was a significant relationship between nurse specialty certification and nurse staffing levels. As the proportion of nurses with a specialty certification increased, both the total number of RN hours per patient day (*r_s_* = .434, *n* = 293, *p* < .001), and the RN-to-patient ratio (r_s_ = .506, *n* = 293, *p* < .001) increased.

### Mediator Effects & Moderator Effects

There were no significant indirect effects of medical treatments on GC, indicating that nurse certification did not mediate the relationship between medical treatments and GC (Table 2). However, nurse certification moderated the effects of the insulin regimen administered and nutritional treatments on GC (Table 3). The interaction between nurse certification and type of insulin regimen administered, accounted for a significant amount of the variation in mean blood glucose levels during hospitalization (Δ*R^2^* = .011, *F*[2, 287] = 3.797, *p* = .024). Among patients who were administered basal-bolus insulin, a 1-level (low-to-medium, medium-to-high) increase in the percentage of specialty certified nurses on the unit, decreased mean blood glucose by an average of 15.7 mg/dl (*p* = .007); however, among those who were administered sliding scale insulin, a 1-level of increase in the percentage of specialty certified nurses on the unit had no effect on mean blood glucose (*p* = .250) [Figure 1]. The interaction between nutritional feedings (enteral or parenteral) and nurse certification also accounted for a significant amount of the variation in mean blood glucose during hospitalization (Δ*R^2^* = .008, *F*[3, 289] = 4.209, *p* = .041). For each level of increase in the percentage of specialty certified nurses on the unit, the difference in mean blood glucose between those who received nutritional feedings and those who did not, decreased by an average of 37.3 mg/dl (*p* = .041); however, among patients who did not receive nutritional feedings, a 1-level change in the percentage of specialty certified nurses on the unit did not affect mean blood glucose (*p* = .449) [Figure 2]. Nurse certification did not moderate the effects of any medical treatment nor nutritional feedings on the percentage of dysglycemic blood glucose testing results, nor on glycemic variability.

**Table 2.** Percentage of Specialty Certified Nurses as a Mediator of the Effects of Medical Treatments on Glycemic Control (*N* = 293).

| **Predictor (X)** | **Indirect Effects of X on *AVGGLU*<sup>†</sup>** |  |  |
| --- | --- | --- | --- |
|  | ***b*** | ***SE*** | ***95% C.I.*** |
| Glucocorticoids | -.026 | .036 | -.1399, .0135 |
| Beta-blockers | -.002 | .021 | -.0728, .0270 |
| Oral antidiabetes medications | -.011 | .047 | -.1460, .0509 |
| Insulin regimen | -.013 | .018 | -.0598, .0114 |
| Feedings | .016 | .055 | -.0474, .1876 |

| **Predictor (X)** | **Indirect Effects of X on *PGLU*<sup>††</sup>** |  |  |
| --- | --- | --- | --- |
|  | ***b*** | ***SE*** | ***95% C.I.*** |
| Glucocorticoids | -.012 | .018 | -.0758, .0088 |
| Beta-blockers | -.001 | .010 | -.0340, .0148 |
| Oral antidiabetes medications | -.006 | .028 | -.1247, .0254 |
| Insulin regimen | -.006 | .009 | -.0347, .0055 |
| Feedings | .008 | .027 | -.0237, .1083 |

| **Predictor (X)** | **Indirect Effects of X on *GV*** |  |  |
| --- | --- | --- | --- |
|  | ***b*** | ***SE*** | ***95% C.I.*** |
| Glucocorticoids | -.005 | .027 | -.0972, .0291 |
| Beta-blockers | .000 | .015 | -.0321, .0303 |
| Oral antidiabetes medications | -.001 | .032 | -.0856, .0505 |
| Insulin regimen | -.004 | .012 | -.0481, .0093 |
| Feedings | .001 | .040 | -.0659, .0981 |

\*Significant models at $p < .05$ . <sup>†</sup>*AVGGLU* = Mean blood glucose level;
<sup>††</sup>*PGLU* = Percentage of dysglycemic blood glucose testing results;
*GV* = Glycemic variability.

**Table 3.**
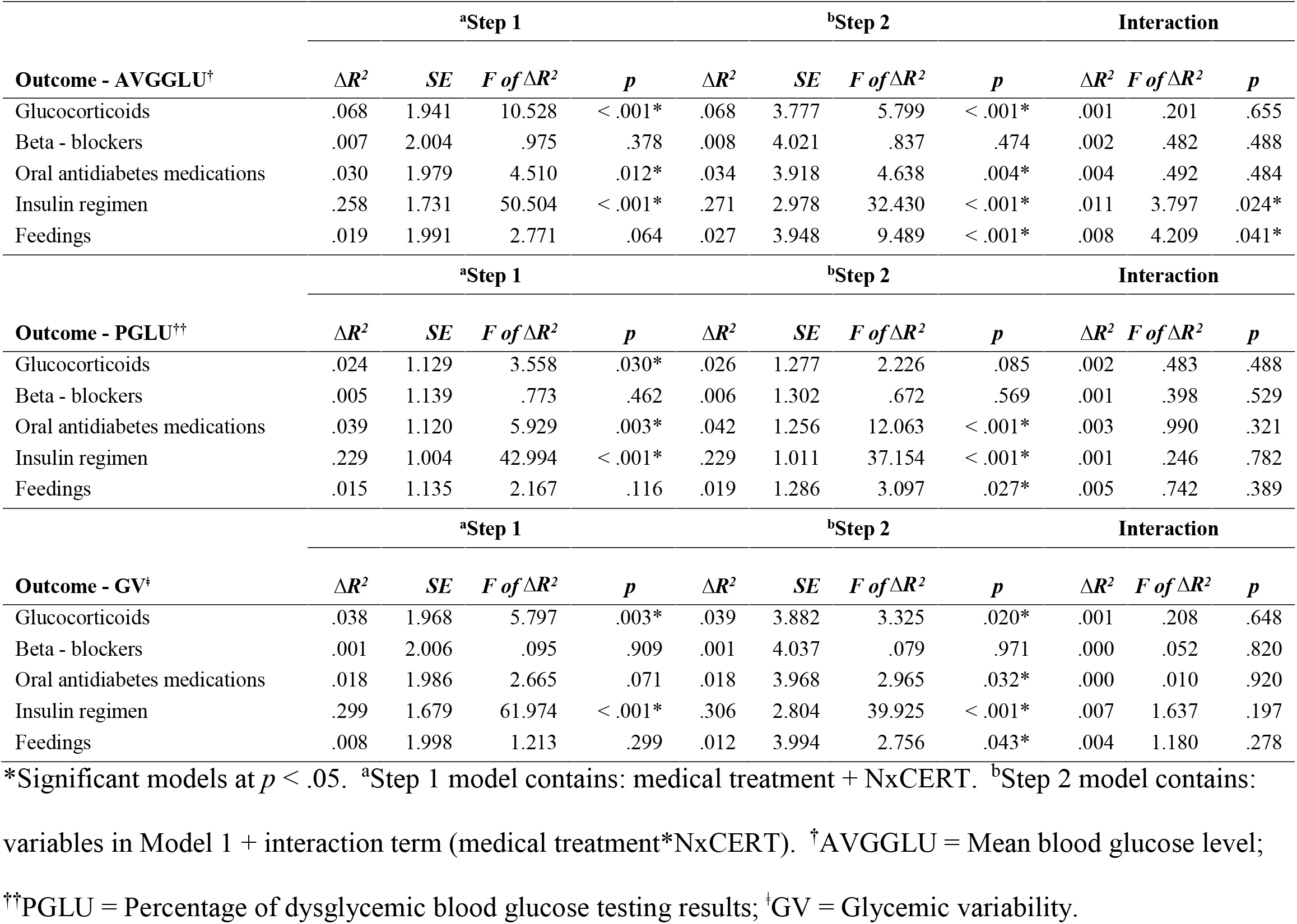
Summary of Hierarchical Multiple Regression Models: Percentage of Specialty Certified Nurses as a Moderator of the Effects of Medical Treatments on Glycemic Control (*N* = 293).

**Figure 1.**
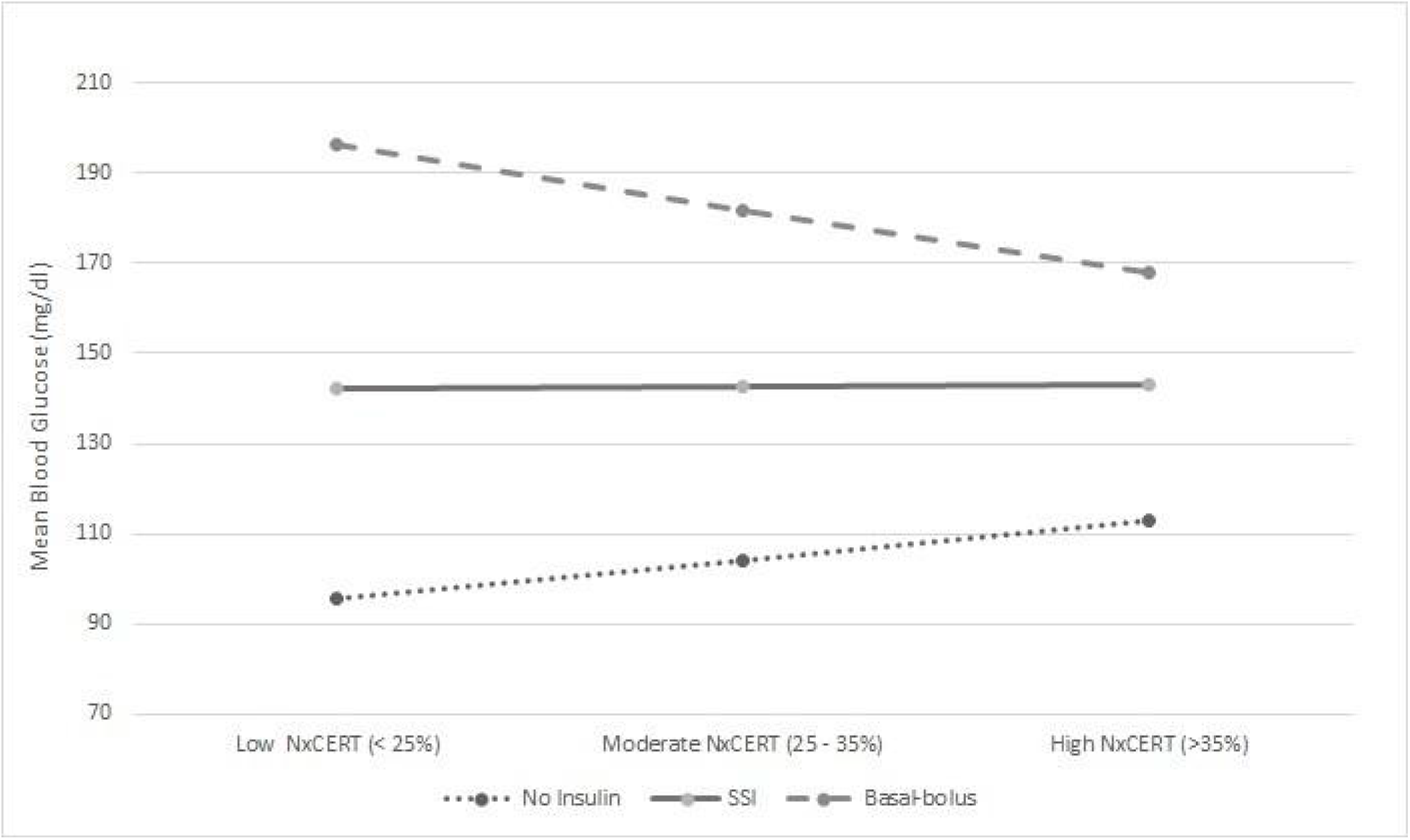
Interaction plot for the effects of the type of insulin regimen administered during hospitalization and the percentage of nurses on the unit with a specialty certification on mean blood glucose levels during the period of hospitalization. *Note*. Among patients who received basal-bolus therapy, each level of increase in the percentage of specialty certified nurses on the unit (NxCERT) was associated with an average decrease in mean blood glucose levels of 15.7 mg/dl. No effect was found among those who received therapy using sliding-scale insulin protocols (SSI).

**Figure 2.**
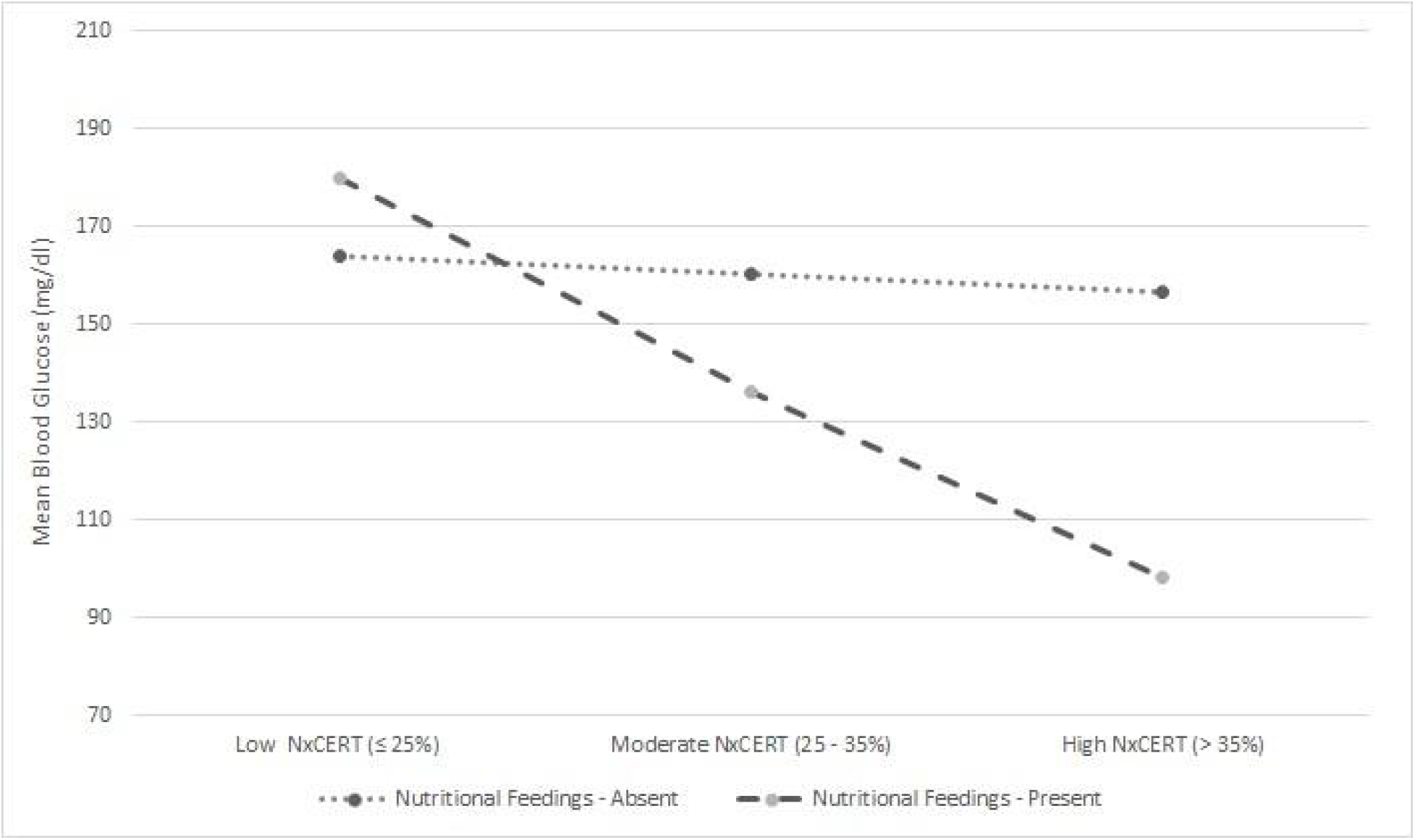
Interaction plot for the effects of nutritional feedings and the percentage of nurses on the unit with a specialty certification on mean blood glucose levels during the period of hospitalization. *Note*. Among patients who received nutritional feedings, each level of increase in the percentage of specialty certified nurses on the unit (NxCERT) was associated with an average decrease of 37.3 mg/dl in mean blood glucose.

## Discussion

While this study did not uncover any direct causal link between the nurse certification and GC, the derivation of causal inferences in mediation modeling is not based solely on the results of statistical testing but rather, evidence of causality should also be evaluated according to accepted practice and guidelines (Hayes, 2013). In the context of the hospital environment, the achievement and maintenance of GC presents many challenges because it is partially driven by the timely performance of routines or “processes” of care surrounding patient nutrition, blood glucose monitoring and insulin administration (Draznin et al., 2013). Nurses deliver many of the care processes required in the management of diabetes within hospitals and are assumed to be key members of multidisciplinary teams caring for hospitalized patients with diabetes (McHugh et al., 2011; Moghissi et al., 2009). Clearly, the theoretical basis for asserting that the activities of certified nurses might exert some direct causal effects on the achievement of inpatient GC remains. Follow-up studies should involve more complex modeling methodologies to help determine the consistency of these findings and to ascertain the possibility of false-positive results. Additional efforts should be directed towards the identification of diabetes-specific nursing care processes within inpatient settings. Without such research we will not be able to identify deficiencies and strengths in current practice, barriers and facilitators of quality diabetes-specific care at the bedside, or allow for the design of efficient and effective nursing interventions that lead to improvements in GC outcomes within hospital settings. The findings of this study did corroborate those of prior research supporting the relationship between medical treatments and inpatient GC. With the exception of beta-adrenergic blockers, most of the medical treatments examined (glucocorticoids, oral antidiabetes medications, type of insulin regimen, and nutritional feedings) were directly linked to changes in GC during hospitalization. Those treatments involve physiological mechanisms known to effect GC and exhibit temporal antecedence to changes in inpatient GC, as such, their role as causal factors in the achievement and maintenance of inpatient GC cannot be overlooked. Further, despite evidence-based recommendations from professional organizations discouraging the use of sliding scale insulin and supporting the use of basal-bolus insulin regimens during hospitalization (ADA, 2015; ADA, 2020), a substantial portion of the patients in this study had their diabetes managed using sliding scale insulin regimens. Thus, among those who received sliding-scale insulin, increasing proportions of certified nurses on the unit had no effect; however, GC improved with increasing proportions of certified nurses among those who received basal-bolus insulin as well as among those who received nutritional feedings.

The percentage of specialty certified nurses on the unit interacted with the administration of basal-bolus insulin resulting in improvements in GC. Among the 57.3% of patients who received basal-bolus insulin during hospitalization, every level of increase in the percentage of certified nurses on the unit was associated with a significant reduction in mean blood glucose levels. Patients who received basal-bolus insulin had significantly lower mean blood glucose levels, a smaller proportion of dysglycemic blood glucose testing results, and lower glycemic variability than those who received sliding-scale insulin or no insulin at all. Prior studies have linked the use of basal-bolus insulin regimens to lower blood glucose levels (Farrokhi et al; 2012; Korytkowski et al., 2009; Maynard, Huynh, & Renvall, 2008; Schnipper, Barsky, Shaykevich, Fitzmaurice, & Pendergrass, 2006; Umpierrez et al., 2011) and with increased rates of hypoglycemia during hospitalization (Maynard, Huynh, & Renvall, 2008; Umpierrez et al., 2011). The observed increases in the rates of hypoglycemia associated with basal-bolus insulin in prior studies may be partially attributed to the fact that none of the studies accounted for the potential impact of nursing care on inpatient glycemic control. There is evidence suggesting that such a mechanism requires further exploration, indeed the study by Schnipper and colleagues (2006) found that specific medical teams and nursing units were associated with significant decreases in mean blood glucose levels. It is noteworthy that in the current study increases in the proportion of certified nurses on the unit resulted in significant improvements in the average blood glucose level of those who received basal-bolus insulin, but rendered no effect on those who received sliding-scale insulin or no insulin at all during hospitalization. This finding suggests that the benefits of the care delivered by certified nurses to inpatients with Diabetes mellitus are substantial, but that ability for patients to reap full benefits from their care was at least partially dependent upon the administration of evidenced-based glycemic management using basal-bolus insulin regimens.

The administration of enteral or parenteral nutrition increases the risk for hypoglycemia due to sudden interruptions in administration, while failure to promptly adjust antidiabetes medications upon initiation of nutritional feedings may lead to persistent hyperglycemia and poor patient outcomes (ADA, 2015; Pasquel et al., 2010). Thus, in order to maintain GC during hospitalization, experts recommend more frequent monitoring of blood glucose levels among non-critically ill patients receiving nutritional feedings (Umpierrez et al., 2012). In this study, the findings indicate that when models were adjusted for the effects of nurse certification, patients with lower blood glucose levels on average were more likely to receive nutritional feedings during hospitalization compared to those with higher blood glucose levels. However, this relationship was modified due to a significant interaction between nutritional feedings and nurse certification. At the lowest percentage of specialty certified nurses on the unit, patients who received nutritional feedings had mean blood glucose levels that were slightly higher than patients who did not receive nutritional feedings. However, when the percentage of certified nurses on the unit was increased to moderate levels the trend was reversed, patients who received enteral or parenteral feedings now had lower mean blood glucose levels than those who did not. This effect continued, with further reductions observed when the percentage of specialty certified nurses were highest.

The findings of this study indicate that the nutritional status and the medical treatments administered during hospitalization escalate the complexity of providing nursing care to hospitalized patients with diabetes. Adding to that complexity is that fact that patients frequently enter the hospital with a variety chronic medical problems. In this study 73.7% of patients included had been diagnosed with 1 or more diabetes-related complication and another 73.4% of patients suffered from at least 1 additional chronic condition. While this study did not investigate how certified nurses may have contributed to the observed improvements in GC, the Medical-Surgical Nursing Certification Board (2017) dedicates 14% - 16% of the questions on the certification exam to the care of patients with diabetes and other endocrine disorders, ensuring a minimum level of competency. These findings suggest that specialty certified nurses may be better prepared to manage the complexities of inpatient diabetes care.

Lastly, when comparing the relationship of nurse certification to nurse staffing indicators, the results were contradictory. As the percentage of certified nurses on the unit increased, the total RN hours per patient day increased. It is possible that an interaction effect between total RN hours per patient day and the percentage of certified nurses contributed to improvements in GC. In 2011, McHugh and colleagues found that in non-teaching hospitals, increases in nurse staffing levels were associated with decreases in the occurrence of diabetic emergencies. Thus, nurse staffing levels may have some significant independent effects on GC. However, as RN-to-patient ratios increased, so did the percentage of certified nurses on the unit. These contradictory findings suggest that the association between indicators of nursing care quality and GC is more complex than could be examined within the confines of this study and may have been influenced by unmeasured factors such as teamwork (Kalisch & Lee, 2010).

### Relevance to Clinical Practice

Professional organizations such as the ADA (2015), the American Association of Diabetes Educators (AADE) (2016), and the Endocrine Society (Umpierrez et al., 2012) have adopted practice recommendations that highlight the period of hospitalization as an opportunity to engage patients in diabetes-specific self-management education and to offer referral for further education and follow-up in the community. Given the complexity of diabetes, the AADE (2016) has recommended that such education should be delivered by an interdisciplinary team that includes a clinician who is certified as a diabetes educator. Only 18% of certified diabetes educators are employed in hospital settings (AADE, 2016), thus access to expert diabetes educators during hospitalization may be improbable for many patients. In light of the disproportionate representation of persons with diabetes within hospital settings and the economic pressures to reduce hospital lengths of stay and penalties attached to patient readmissions in some countries, encouraging more staff nurses to become specialty certified may prove invaluable towards the achievement high-quality inpatient diabetes care.

### Strengths

This study used a substantial sample size of 293 patient records to examine the relationship between nurse certification, medical treatments, and GC. To better approximate conditions at the bedside, the study used patient-specific variations in exposure to higher or lower levels of certified nurses on the unit to examine their impact on GC. In addition, this current study did not exclude diabetic patients with specific primary or secondary diagnoses, reflecting the realities of the hospital setting and strengthening the external validity of the findings. Lastly, a major strength of this study was the use of mediation and moderation analysis to describe not only whether a relationship existed between nurse certification and GC but also the nature of that relationship and circumstances under which such a relationship might exist.

### Limitations

The study took place at a single not-for-profit community hospital in the United States that was on the journey to recognition for nursing excellence during the time of the study, thus levels of certification found at the study site may not be typical of community hospitals. Few patient records included in this study were from patients who had been administered nutritional feedings, this may have reduced the power and precision of the analysis. As with all studies involving retrospective data, the accuracy of the findings rely on the consistency, accuracy and completeness of documentation. Administrative data on nurse staffing levels were based on the midnight census and may not have captured variations in staffing throughout the day. Lastly, the hospital setting is complex and the number of factors that might be conceptualized as contributing to GC are extensive. Despite careful selection of the variables included in this study, the possibility that some of the findings may have been confounded by the presence of latent or unmeasured variables cannot be dismissed.

## Conclusions

This study provided preliminary evidence that specialty certified staff nurses contributed to improvements in GC among subgroups of non-critically ill patients who were administered basal-bolus insulin or nutritional feedings during hospitalization. The findings suggest that patients who received a basal-bolus insulin regimen for the treatment of their diabetes during hospitalization and those patients who were prescribed and administered nutritional feedings drew added benefits when nursing care was delivered during times in which the relative proportion of certified nurses on the unit were higher.

## Data Availability

The data set analysed during the study is not publicly available but is available from the corresponding author on reasonable request.

